# Work-related and Personal Predictors of COVID-19 Transmission

**DOI:** 10.1101/2020.07.13.20152819

**Authors:** Paul Anand, Heidi Allen, Robert Ferrer, Natalie Gold, Rolando Manuel Gonzales Martinez, Evan Kontopantelis, Melanie Krause, Francis Vergunst

## Abstract

The paper provides new evidence from a survey of 2000 individuals in the US and UK related to predictors of Covid-19 transmission. Specifically, it investigates work and personal predictors of transmission experience reported by respondents using regression models to better understand possible transmission pathways and mechanisms in the community. Three themes emerge from the analysis. Firstly, transport roles and travelling practices are significant predictors of infection. Secondly, evidence from the US especially shows union membership, consultation over safety measures and the need to use public transport to get to work are also significant predictors. This is interpreted as evidence of the role of deprivation and of reactive workplace consultations. Thirdly and finally, there is some, often weaker, evidence that income, car-owership, use of a shared kitchen, university degree type, risk-aversion, extraversion and height are predictors of transmission. The comparative nature of the evidence indicates that the less uniformly stringent nature of the US lockdown provides more information about both structural and individual factors that predict transmission. The evidence about height is discussed in the context of the aerosol transmission debate. The paper concludes that both structural and individual factors must be taken into account when predicting transmission or designing effective public health measures and messages to prevent or contain transmission.

**JEL Codes:** I1 I12 I14 I18

## 1. Introduction

Potentially, preventing the transmission of COVID-19 related to work and amongst the poor, saves lives while contributing to other economic and social priorities. A large amount of scientific research has focussed on patterns of spread and underlying mechanisms of transmission but as economies and societies reopen, it is important to know more about the role of workplace and personal factors as predictors of transmission.[1-2] Heightened risks implied by spatial patterns [3] and attached to certain work-roles have emerged as important but there are many aspects of employment and consumption activities that are likely to contribute to transmission that have barely been researched. In addition, and closely connected, there is a growing body of knowledge about personal factors that contributes to mortality but (with the exception perhaps of ethnicity) only a smaller amount of literature of personal traits and circumstances relating to transmission risk within community settings.[4-5].

To limit the spread of the virus, it is therefore important to study work-related and personal factors that contribute to or could limit the spread of the virus. This paper therefore reports on the development of data relating to a new set of diverse workplace and personal factors. More specifically, using a survey of 2000 working age adults in the US and UK, the paper estimates regression models in which work, personal factors and a range of demographic controls are used to predict experience of Covid-19. Both countries are examples of high-income market economies are distinct from others in two ways. Unlike some Asian countries, they do not have recent similar infection-spread experiences on which to draw and unlike many European countries, they do not have civil law traditions based on a ‘strong’ conception of the state. Yet the US and UK differ in the extent and manner in they provide access to health care and welfare support. Furthermore, the US has experienced a lockdown that has varied significantly between states.

For these two countries, the paper draws on a new database related to estimate regression models of transmission experience. The dataset contains several variables related to transmission experience while the analysis focuses on the possession of a medical diagnosis or positive test self-reported by the respondent. Analysis for the US provides evidence of infection risk related to transport related employment, working with reduced earnings, belonging to a union, workplace consultation about safety measures, being on a zero hours contract and having to use public transport to go to work are significant predictors. At the personal level, controlling for race and age, being in the lower income groups, having a shared kitchen, a quantitative university degree, using cash to pay and car ownership are also significant predictors of infection diagnosis and positive tests. (There is also some evidence in pooled univariate analysis that the probability of infection is related to other variables including risk preference and extraversion.) Results for the UK are less statistically significant but generally similar qualitatively probably due to the more uniform nature of its lockdown.

Three emerging themes for public health, individual behaviour and research are discussed. (i) *Transport* as an employment setting or mechanism for getting to work is a predictor of transmission and so safety in such contexts should be prioritised. (ii) Features of the *workplace and employment* are also significant predictors of transmission within the community and there is therefore a need for a much fuller understanding of work and commuting measures so that public health and economic perspectives and priorities can be more fully integrated. (iii) A diverse range of *personal attributes and circumstances* predict infection and relate both to behavioural and distributive issues. In some cases, the predictors speak to issues of deprivation and poverty. In one case, the impact of height, even after controlling for income and sex, may also be relevant for the recent debate about droplet and aerosol transmission. In any case, these personal differences should be taken into account when designing public health measures, giving medical advice and designing labour market policies.

The rest of the paper is structured as follows. Section 2 summarises the key variables and statistical techniques used. Section 3 carries the main results while section 4 discusses these results in community and policy contexts, some limitations and possibilities for follow-up work.

## 2. Methods and Materials

### Data

The database described below (and in the online materials) from which the variables are drawn was developed during a period when general scientific pathways of transmission were becoming more widely accepted but there was little evidence on some of the possible predictors and mechanisms in US and UK communities. It provides a mix of standard and novel data on a range of personal, work, home and community factors.

### Personal Factors

This paper draws on personal variables related to risk-aversion, personality, university degree, car ownership, sex, household income and the use of cash to make payments. Ethnicity and age are also used as controls. See Table 1.

**Table 1.**
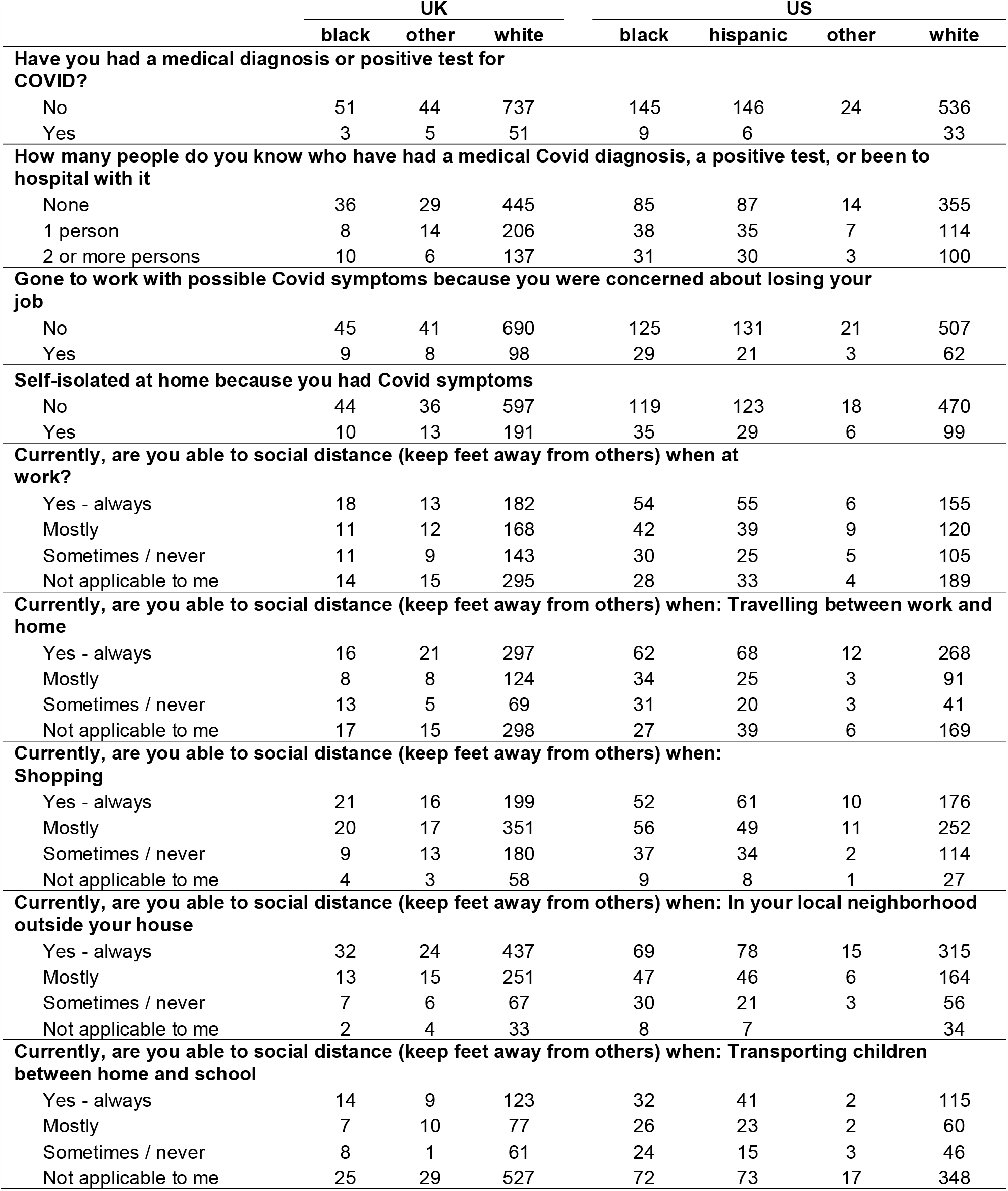
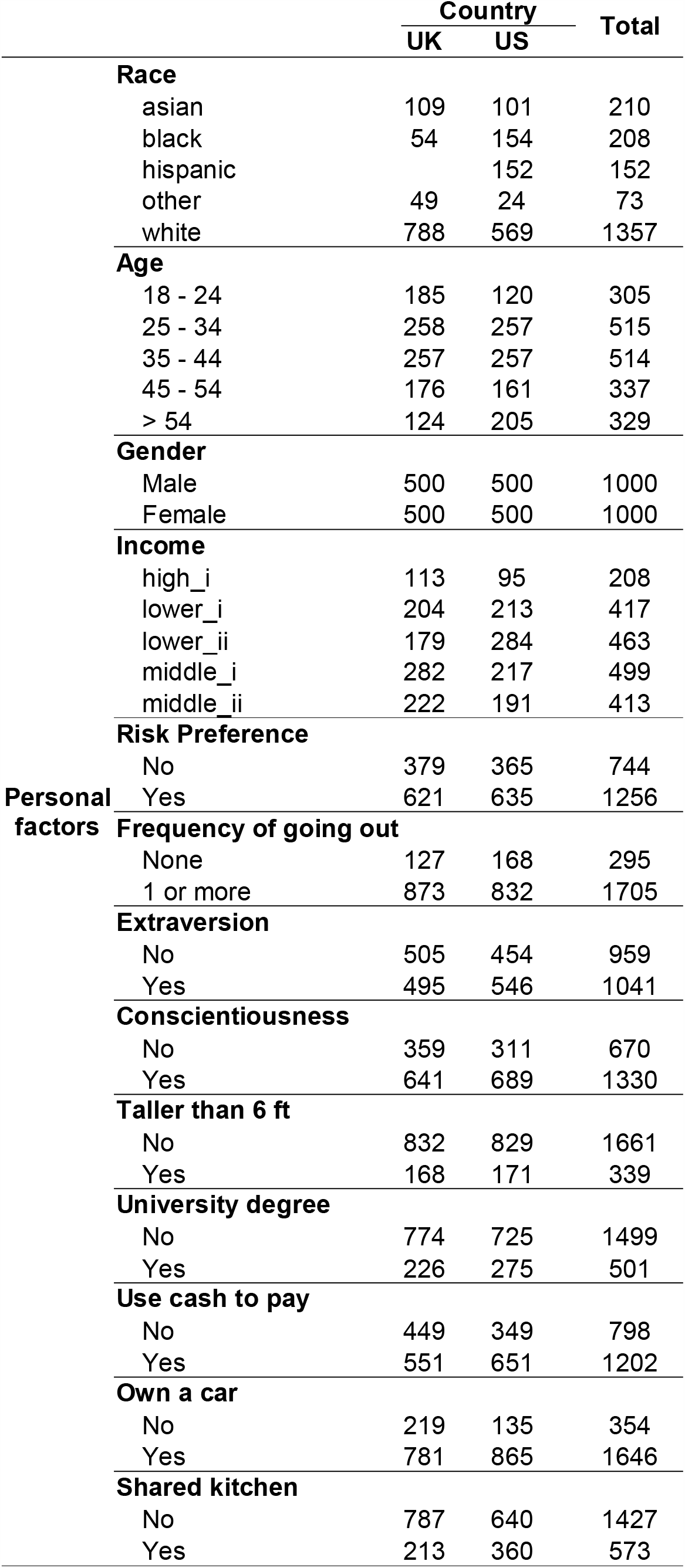
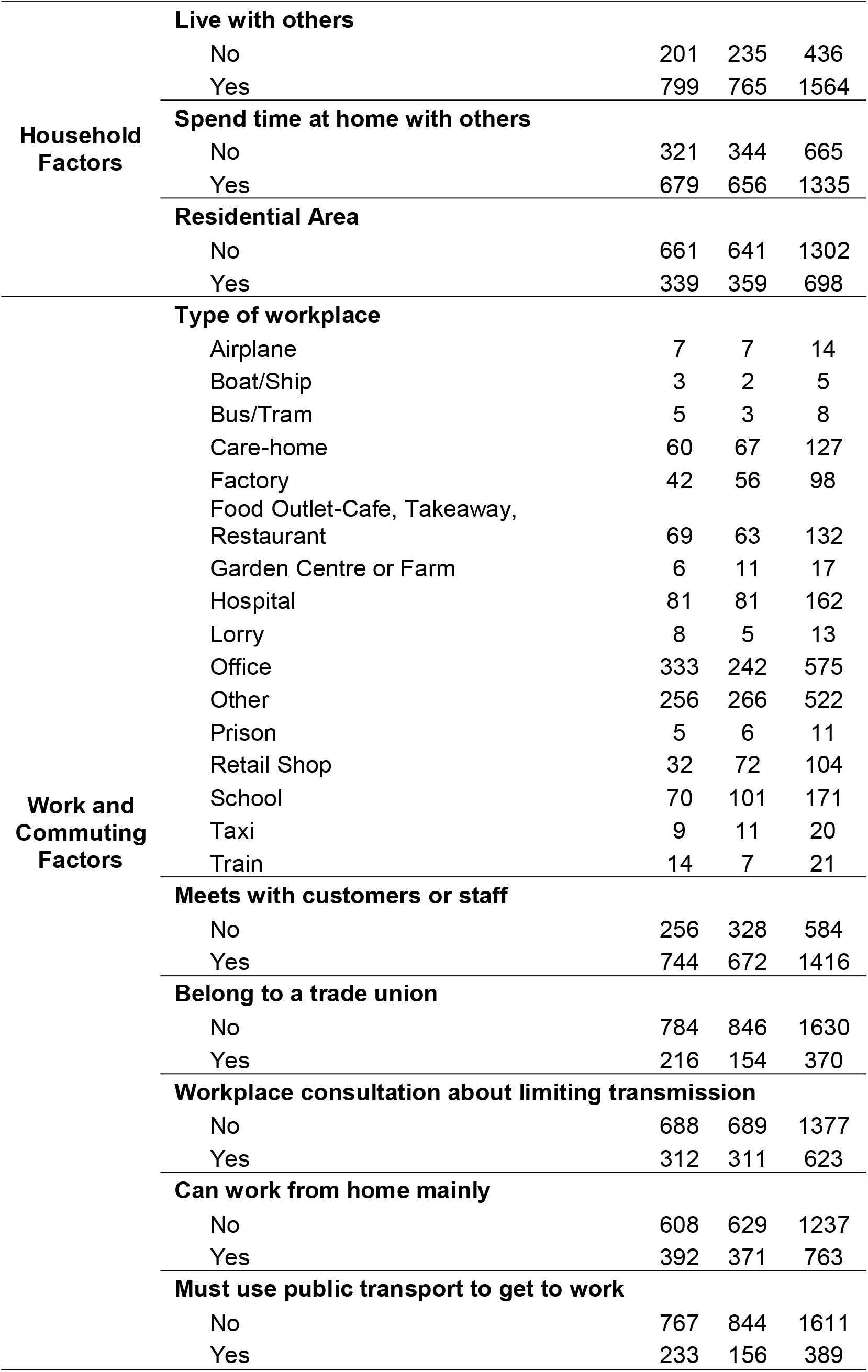
Descriptive Statistics for the UK and US

To assess risk aversion, the survey contains a single question that has been used previously and validated against other measures in economics.[6] Risk preference plays a central role in the theorising of economic behaviour and it is reasonable to hypothesise that it also plays an important role in transmission related behaviours. In addition, the database (see online materials) includes information on personality, already connected to information and attitudes about risk-taking,[7] and it was hypothesised that extraversion could also predict of transmission. Extraverts are more likely than others to engage in social activity and the trait was measured using two questions reverse coded from a widely used short form personality battery.[8] Height has been associated both with health [9] and income and so the database assesses also tallness which could potentially be a protective or a risk factor depending inter alia on transmission patterns. If large downward falling droplets were more significant then taller people might be expected to be less at risk. However, it has recently been argued that aerosol (fine particle) transmission, important for influenza [10], may also be important for the transmission of Covid-19 [11-12]. If overhead airflows did play a role early then, then taller people might be at greater risk of infection. The use of cash to pay was noted as a risk factor early on [10] as paper notes and coins are subject to sequential physical contact and a variable on this is also included in the analysis.

Several other personal predictors are also available and included in the analysis. Sex and ethnicity have been found to be risk factors for mortality [13-14] and may also be connected to transmission. Working patterns or even feeling safe outside of a house alone may, for example, cause some men to undertake activities outside the home and therefore be more at risk of infection. On the other hand, the concentration of women in caring professions may place some groups of women at greater risk. Ethnicity could work in similar ways if some groups are disproportionately found in riskier jobs or more crowded residential areas. For the purposes of this study sex age and ethnicity are used along side other demographic controls discussed below.

Although involvement in lorry driving has also been implicated in the spread of Covid-19,[15] car ownership might also be a significant protective factor if the use of private transport enables individuals and family members to social distance for more of the time. To the extent that safety is a good, household income is likely to be an indicator of a range of omitted factors that impact risk such as living in a tower block or having access to a private garden. Finally, a variable is included that indicates the frequency with which the individual went outside the house prior to March (see online supplement). This is likely to be more relevant for onsets that take place prior to the lockdown period.

### Work Related Factors

A second set of predictors relate to work and commuting. The main workplace setting is recorded in a variable with fifteen response categories. Some of these are already known to contribute to transmission,[16] particularly those related to transport, though less is known about others. In addition, there are two variables that record whether a person is forced to use public transport to commute to work and whether they are able to work mainly from home.[17] Both are risk factors although the sign on the ability to work from home is difficult to assess *a priori*. On the face of it, the ability to work from home enables a person to avoid social interactions at work and when commuting but it could for some be offset by additional risks from household or local community contacts. For example, if working from home is associated with greater use of small local shops where distancing is difficult as might be the case in places like New York or London, then working from home could also be a risk factor for some. At the time of variable development, unions in the UK were being reported in the media for their advocacy of health and safety issues at work and few if any investigations to date have studied the contribution of trades unions. Again there are several ways in which union membership might come to predict transmission. Workplaces with effective union advocates could have fewer cases of Covid-19 though alternatively, unionisation could be an indicator of a workplace where larger group meetings are relatively easy or where high workplace risks incentivise union membership.

### Demographic and Houshold Controls

A third and final set subset of independent variables concerns factors that are home or community related and used here as controls. One set of questions relate to whether a person lives with children, parents or others. Those living alone might be expected to be less at risk particularly during periods when mobility and activity outside the home are limited by government rules. Alternatively, it could be that those who spend a large amount of time with others at home could be at greater risk or that those living with parents take greater care and so experience less infection on average. The data contains a variable on whether a person spends more 90 mins or more of their time with others at home and has been used in analysis. In addition, a variable that indicates whether a person lives alone or not is also constructed. These predictors are used to estimate models of self-reported infection (whether a person had a medical diagnosis or positive test). In addition, results for knowing others with Covid-19 and the ability to social distance are also reported.^1^

The dataset on which these variables draw was developed by a survey that took place over the first week of June 2020. Samples of 1000 adults in the US and UK were obtained from a professional survey company using quota sampling to obtain a national sample broadly representative for those of working age with some oversampling to reflect contrasts of interest. All survey recruitment and completion was done by electronic means (so via phones or personal computers but not face-to-face meetings). Towards the end of the sampling period some of the quotas were relaxed and the final distribution of socio-economic characteristics of the US and UK samples can be found in Figure 1 and the online materials. The company provides, ex post, a set of weights that can be used to construct nationally representative results and these weights are used at various points. Respondents were paid a small amount for completing the survey which took about 5 minutes on average to complete. It is important to reiterate that survey responses are self reports and that said, overall reported infection rates are comparable to those reported elsewhere for the UK [18] and US [19] bearing in mind the predominance of early transmission experience. Those who became ill at points closer in time to the survey were, plausibly, less likely to respond probably because they were still ill. Selected descriptive statistics for each country are presented in Figure 1.

**Figure 1.**
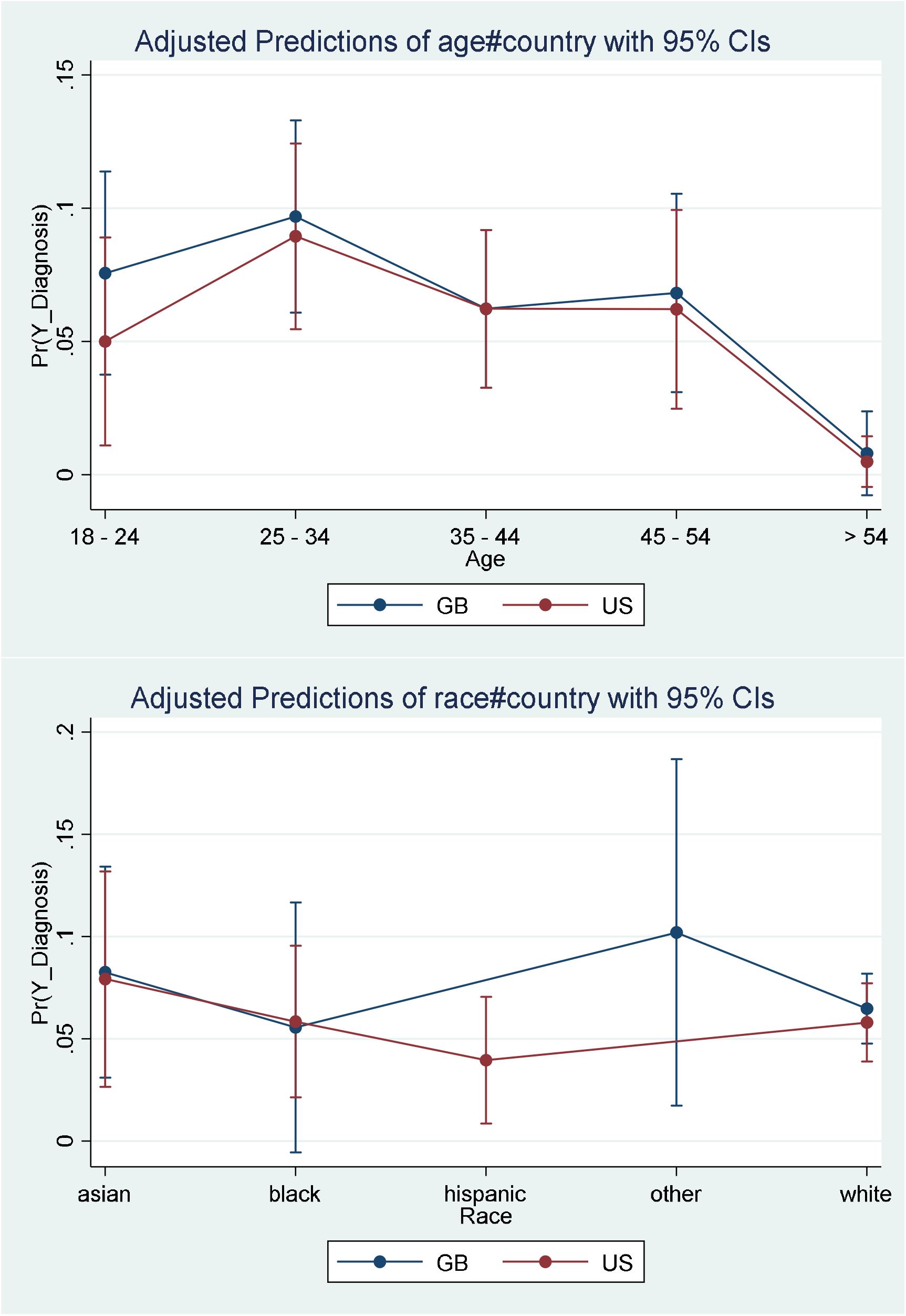
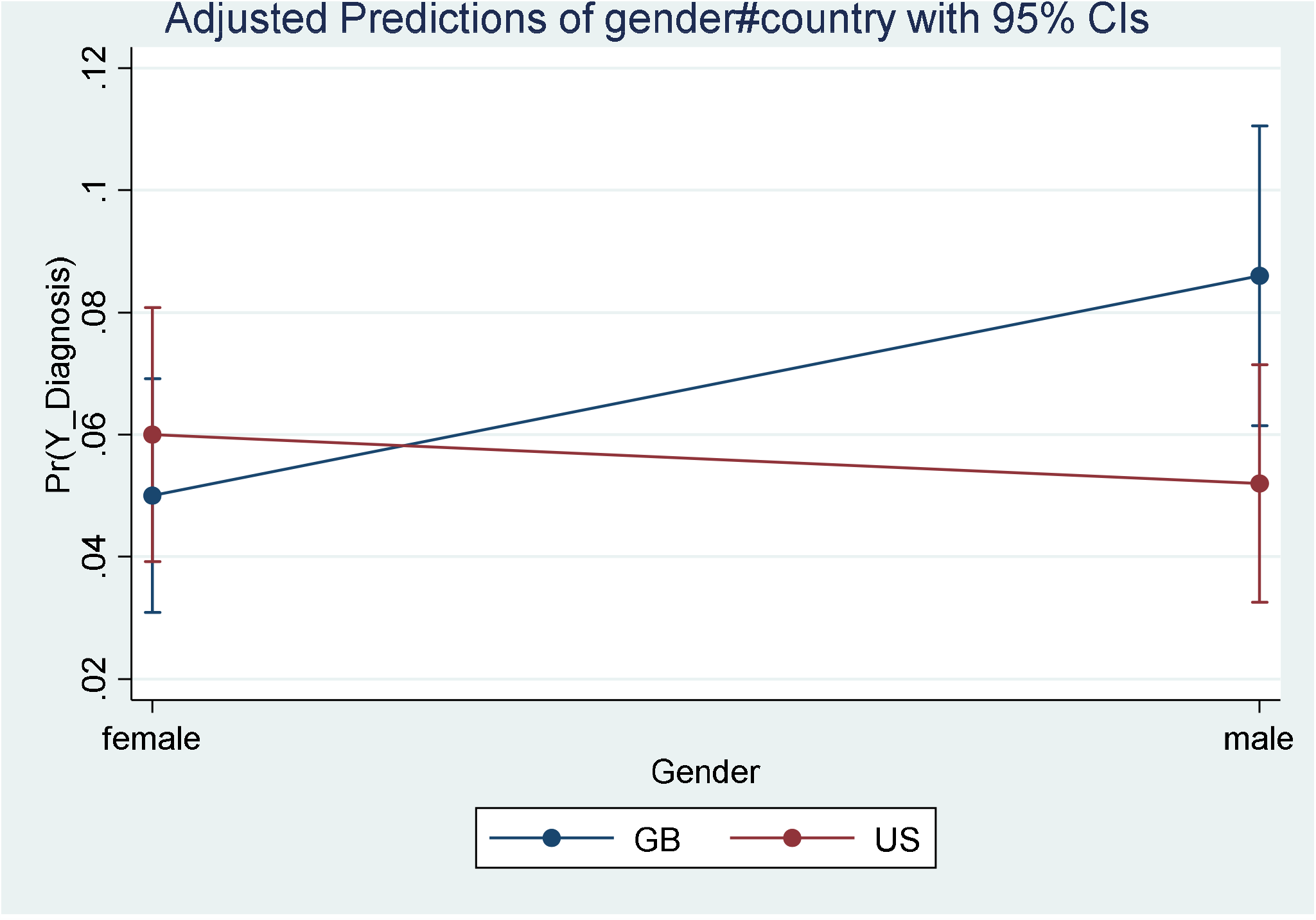

### Methods

The outcome of primary interest was: a) a confirmed diagnosis of COVID-19 (“Have you had a medical diagnosis or positive test for COVID”). In addition, models were estimated for b) knowing someone who was diagnosed with COVID-19 (“How many people do you know who have had a medical COVID diagnosis, a positive test, or been to hospital with it”); and c) having being able to socially distance (“Currently are you able to social distance when at work/ commuting/ shopping/ neighbourhood/ transporting children: Yes-always, Mostly, Sometimes, Never, not applicable”). Separate regression models for the USA and UK samples were estimated, with area of residence modelled as fixed-effects for responders within fourteen states in the US and four constituent countries (England, Scotland, Wales, Northern Ireland or unknown) in the UK.

The exact model applied depends on the type of the outcome variable. For the first outcome, confirmed diagnosis, a logistic model was estimated for a binary dependent variable equal to one for those responders who self-declared to have a positive diagnosis of Covid-19. An additional analysis used sample weights to allow generalisation of the survey result to the population level. The second dependent variable was dichotomized to “none” against “one or more”, and a logistic model was applied to the resulting binary categories, allowing us to estimate the influence of demographic, household and workplace factors on the probability of knowing someone with a positive Covid-19 diagnosis. Without dichotomization, the second dependent variable is a multi-class nominal outcome; hence a multinomial logit was also estimated for the second outcome, using none as the base category for comparison. A Poisson regression model was used as a sensitivity but led to less satisfactory results. Finally, the third outcome variable, which indicates ability to social distance, was calculated with an aggregation over sub-domains: for each question, answers of “Yes-always” were coded as 1, and all other answers were coded to zero; the individual answers were aggregated into a 5-category scale for each respondent. Since the resulting third categorical outcome has an implicit order (ranging from no social distancing at all to complete social distancing or no exposure), an ordered logit model was estimated to evaluate the effects of demographic, household and workplace factors on the cumulative probability of the individuals to keep social distance.

## 4. Empirical Results

In Table 2, results are presented for several models of transmission experience. Variables were selected and categories used based both on clinical or theoretical considerations and univariate analysis (see online materials). Results for the pooled data are similar to those for the US and so these are focussed on in the table. These results suggest that with demographic controls, the probability of transmission depends on a diverse range of work and personal predictors.

**Table 2.**
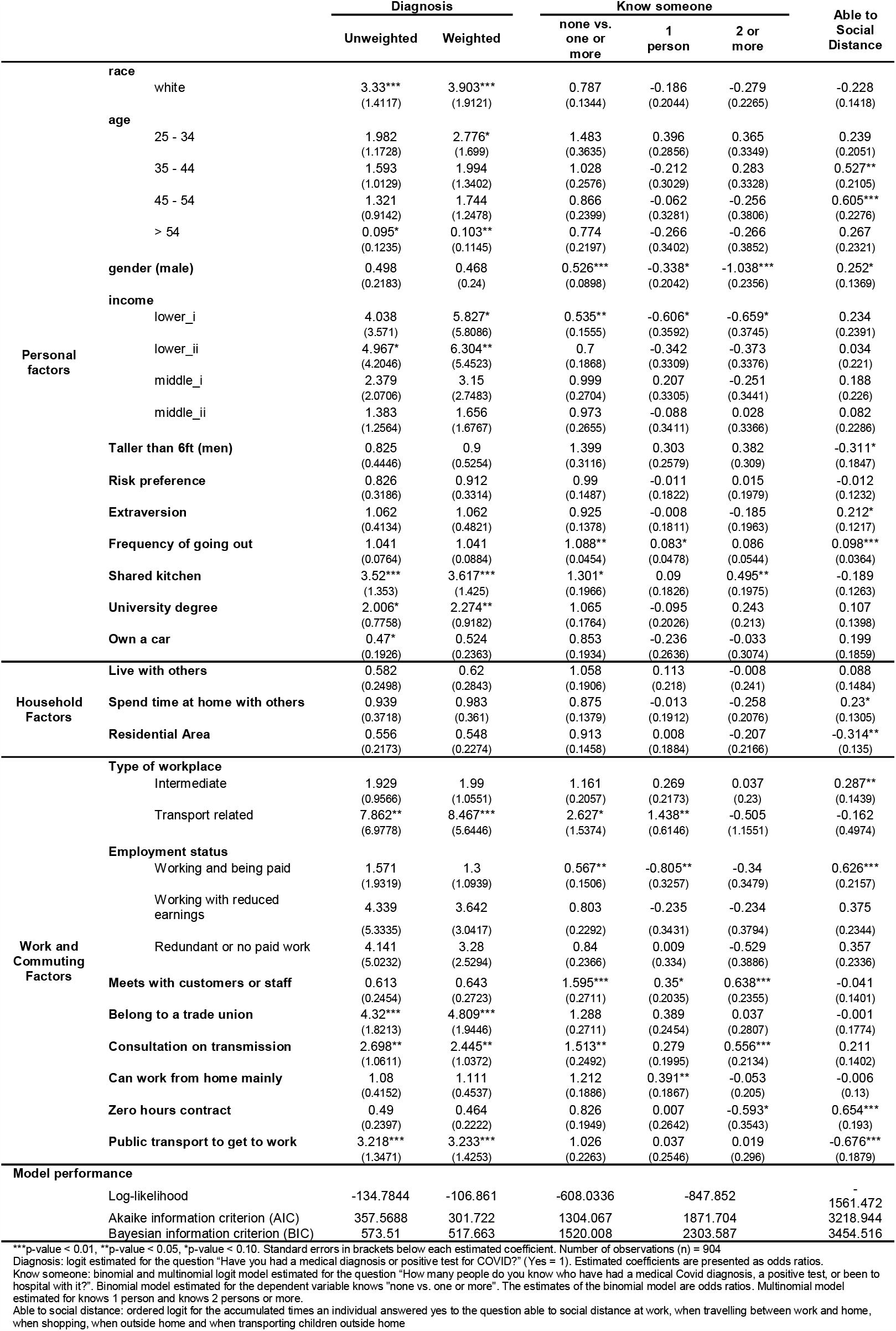
Models of Transmission for the US

Focussing on reported diagnosis and testing, it is evident that several of the work-related variables, with the exception of being able to work from home, are statistically significant. The same is true for some of the personal factors although risk-aversion and extraversion are not as significant as in the univariate analysis. Being in either of the lowest household income groups (ie less than $49,999 pa) is significant whereas none of the controls are. Other aspects of transmission experience exhibit different patterns. For example, knowing others with Covid-19 is significantly related to using a shared kitchen but also to the frequency of having met others at work. The same is true for the ability to social distance – having to go to work on public transport is a risk factor as with transmission but so too is being in the 35-54 age group and in an intermediate risk work-setting.

One of the reasons why results in the UK might differ concerns the more uniform and stringent nature of measures taken. As the purpose of lockdown is to disrupt patterns of transmission that would exist otherwise, it would not be surprising to see weaker associations. To consider the possibility, Table 3 presents similar models for the UK estimated using data excluding those who first experienced symptoms in April and May and therefore were likely to have contracted Covid-19 through social interactions prior to lockdown. These results are presented in Table 3.

**Table 3.**
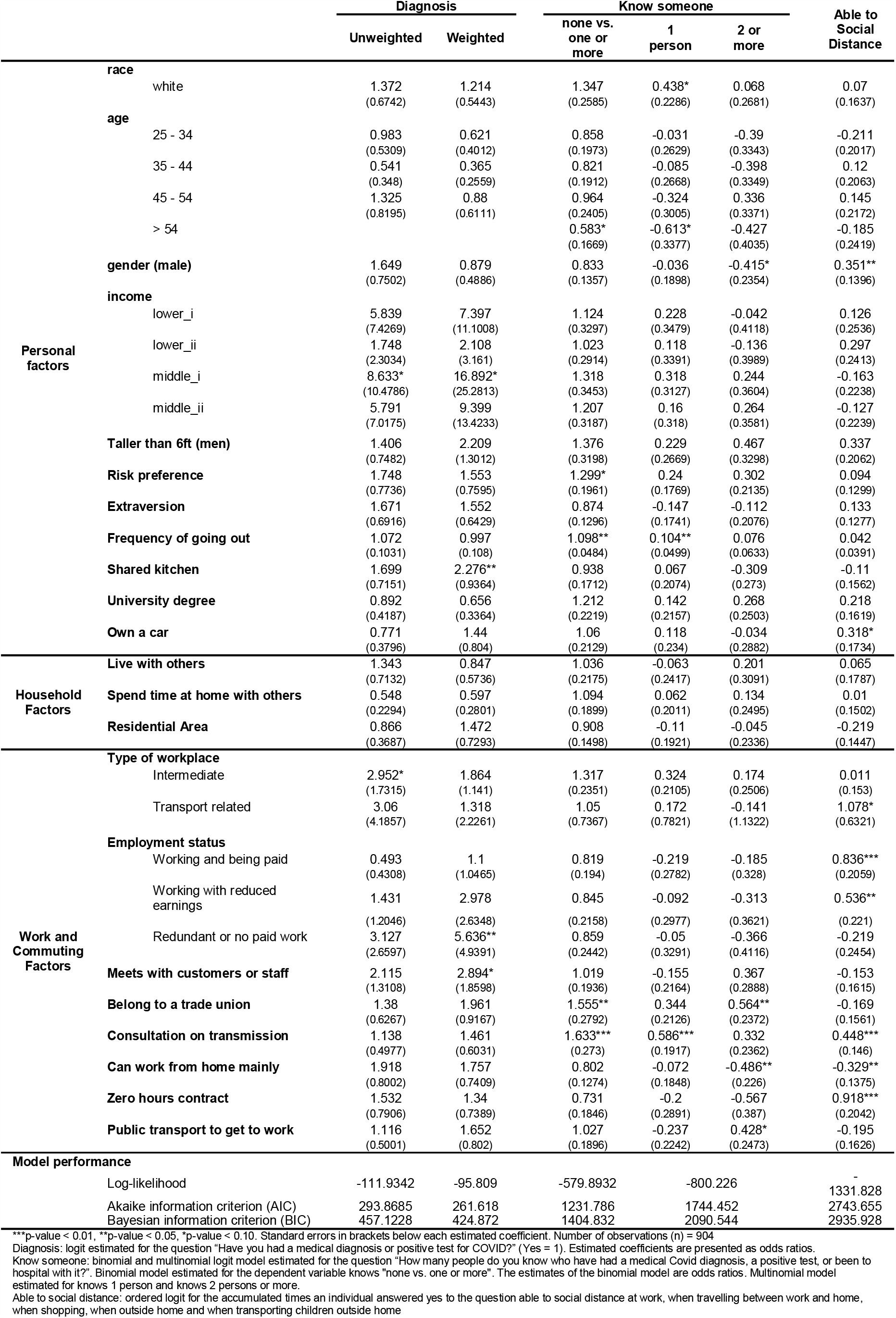
Models of Transmission Experience for UK

It is interesting to note that being male and some types of transport employment are all significant predictors of infection. Most work-related factors are not significant in the UK for diagnosis but some are significant for knowing someone with Covid-19. Owning a car predicts being able to social distance in the UK whereas it does not in the US.

## 4. Discussion

The models of transmission experience add to what is known about transmission in the community[20-21] and confirm some qualitative similarities between transmission predictors in two high income countries. However, the variations in policy response, particularly the greater regional variation in the US lockdown, implies that US experience provides more information about potential pathways for community transmission. A variety of work-related factors are predictive of transmission though sometimes in ways that might not be obvious. Having to go to work on public transport is positively related to transmission in both countries but the effect is considerably stronger in the US. Moreover, union membership is a significant predictor of risk. There are various reasons why this might be the case, as noted above, but the data cannot distinguish between them. In both countries, consultation about transmission reduction measures is positively related to infection and one interpretation is that consultation during the period was more reactive than preventative. The fact that about half of all respondents claim not to have been consulted about such measures is consistent with the interpretation and suggests that safety promotion within the workplace could be monitored and guided by public health and health and safety officials more closely.

Turning to personal factors, it is worth noting that some theoretically supported empirical findings concerning risk aversion and extraversion in the univariate analysis are not significant in the multivariate regression models. Conceptual overlaps as well as correlation in observations may be giving rise to multi-collinearity (though in the main formal VIF tests do not suggest this is a major problem). While not especially worrying from a prediction perspective it is important for public health messaging to take these traits into account as they suggest a need for tailored messaging and interventions. The fact that height is a significant predictor for men suggests that downward droplet transmission may be less important than aerosol transmission (particularly prior to lockdown) in which case the use of specifically designed air purifiers should be further explored. Using a shared kitchen is also a significant factor. While steps have been made in both countries to reduce the use of cash payments, for example, less is known about any guidance or support for those, such as those on low incomes, users of Airbnb, and students who often share kitchens, or other facilities. The practice may be more prevalent than is supposed. It is also worth noting that having a quantitative degree is also not an enabler of social distancing but rather a risk factor for having a diagnosis or positive test. If most respondents held natural science degrees, then it is possible that the natural science background caused respondents to be more willing to seek a medical diagnosis or test. Aversions to testing might derive, therefore, from background interests as well as costs and that too is something that might be factored into the design of test and trace interventions.

While this study adds some novel variables and evidence to the understanding of community transmission within the US and UK, several limitations should be mentioned. In the first place, it would be useful to have larger sample sizes particularly for observations referring substantially to behaviour in a lockdown period. In addition, it would be helpful to have repeated observations so that more could be said about changes over time as well as causality: indeed, it would be useful to have patient or lay input into the development of a fuller set of predictors based on possible causal mechanisms. Furthermore, it was not possible to audit responses. Finally, this study was not designed to engage strongly with the issues of race as they have emerged. The database contains mainly those who report first onset of symptoms early on, possible because those still ill were less inclined to participate in surveys. The higher levels of infections of Whites in the survey is consistent with a pattern of infection in which more affluent population members are exposed first to spread via international sources from Europe and elsewhere, while internal transmission then proceeded more rapidly amongst the poor often at greater risk and less able to take avoidance measures.

These limits aside, the study implicates transport related employment and travel in various ways with transmission risk, identifies novel employment related predictors of infection risk, and provides evidence of ways in which personal traits, circumstances and behaviours impact on transmission experience. This is as far as we know one of it not the first study to investigate a range of work and personal predictors of Covid-19 transmission risk comparatively in the US and UK. If similar work and related activity data were collected routinely along with other medical data, it should be possible to identify types of settings where transmission is most likely to take place. This in turn could help refine the preventative measures taken or advised.

## Data Availability

Data is available via a link in the supplementary materials

## Supplementary Materials

Link to Data and questionnaire from which variables drawn https://osf.io/v9t8a/?view_only=8531e8dd672f41e6bf532e280a2f31e6

Key to per annum household income categories

US

Lower i Under $25000

Lower ii Between $25000 and $49000

Middle i Between $50000 and $74999

Middle ii Between $75000 and $99999

UK

Lower i Under £12500

Lower ii Between £12500 and £18499

Middle i Between £18500 and £49999

Middle ii Between £50000 and £62499

Source: https://resources.pollfish.com/pollfish-school/household-income-mapping/

**Table o1.**
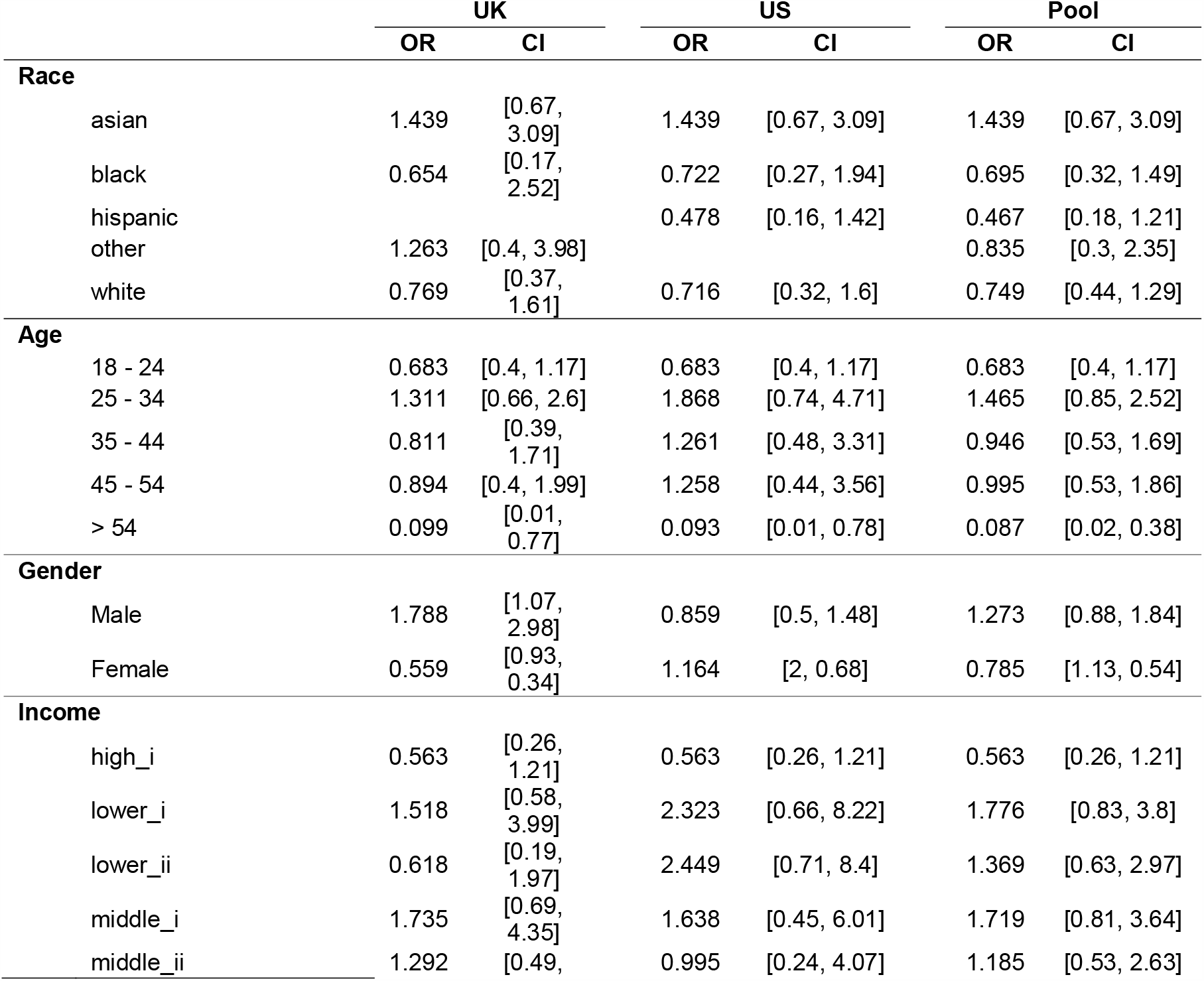
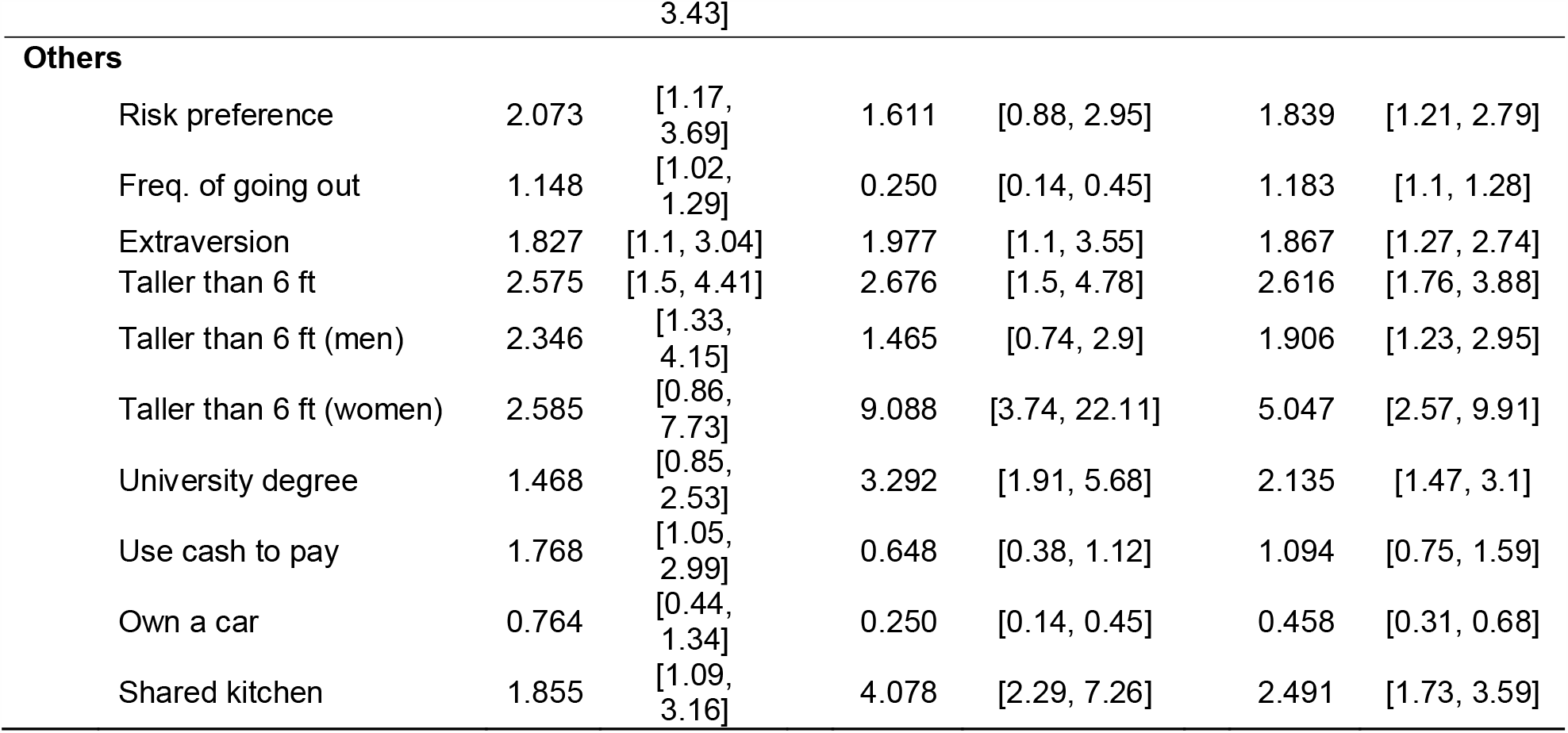
Univariate odds ratios for Personal Factors.

## Univariate odds ratios for Household Factors

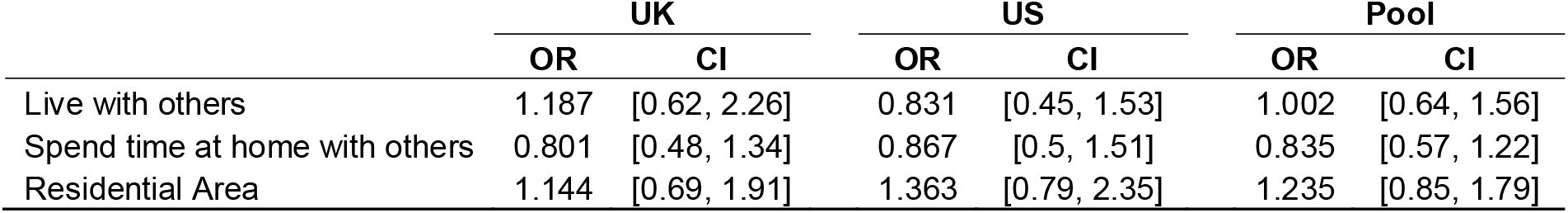

|  | UK |  | US |  | Pool |  |
| --- | --- | --- | --- | --- | --- | --- |
|  | OR | CI | OR | CI | OR | CI |
| Live with others | 1.187 | [0.62, 2.26] | 0.831 | [0.45, 1.53] | 1.002 | [0.64, 1.56] |
| Spend time at home with others | 0.801 | [0.48, 1.34] | 0.867 | [0.5, 1.51] | 0.835 | [0.57, 1.22] |
| Residential Area | 1.144 | [0.69, 1.91] | 1.363 | [0.79, 2.35] | 1.235 | [0.85, 1.79] |

## Univariate odds ratios for Work and Commuting Factors

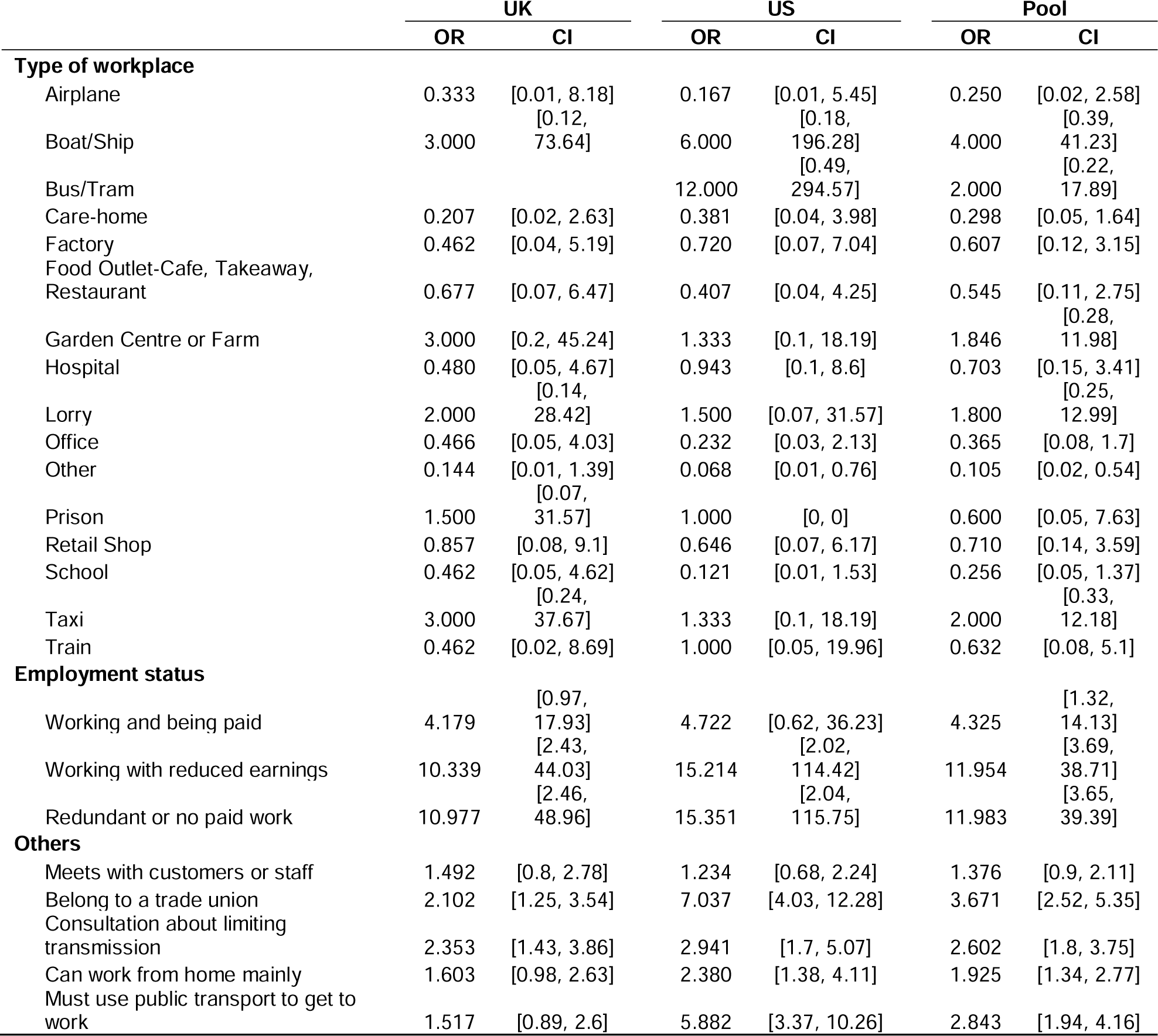
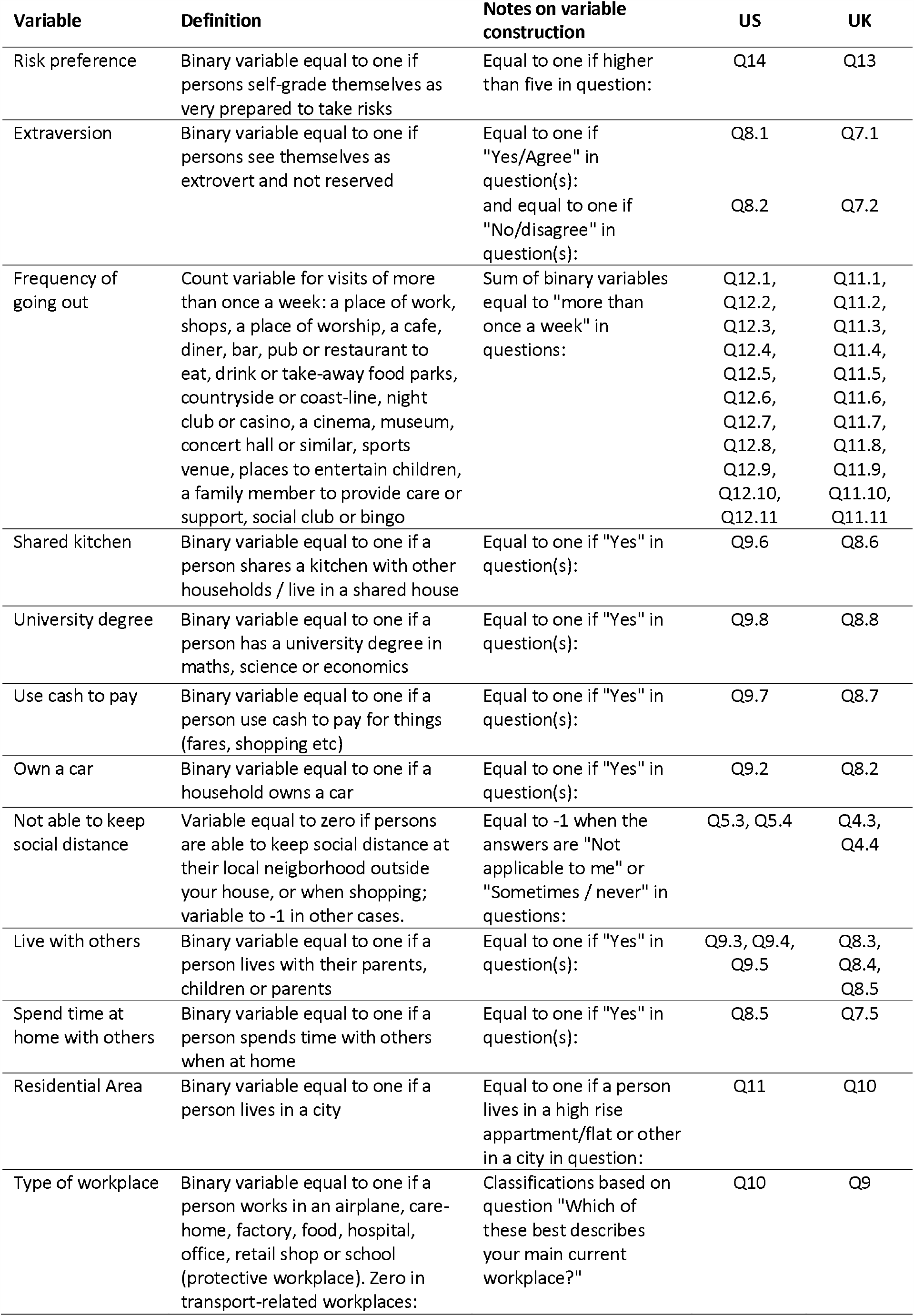
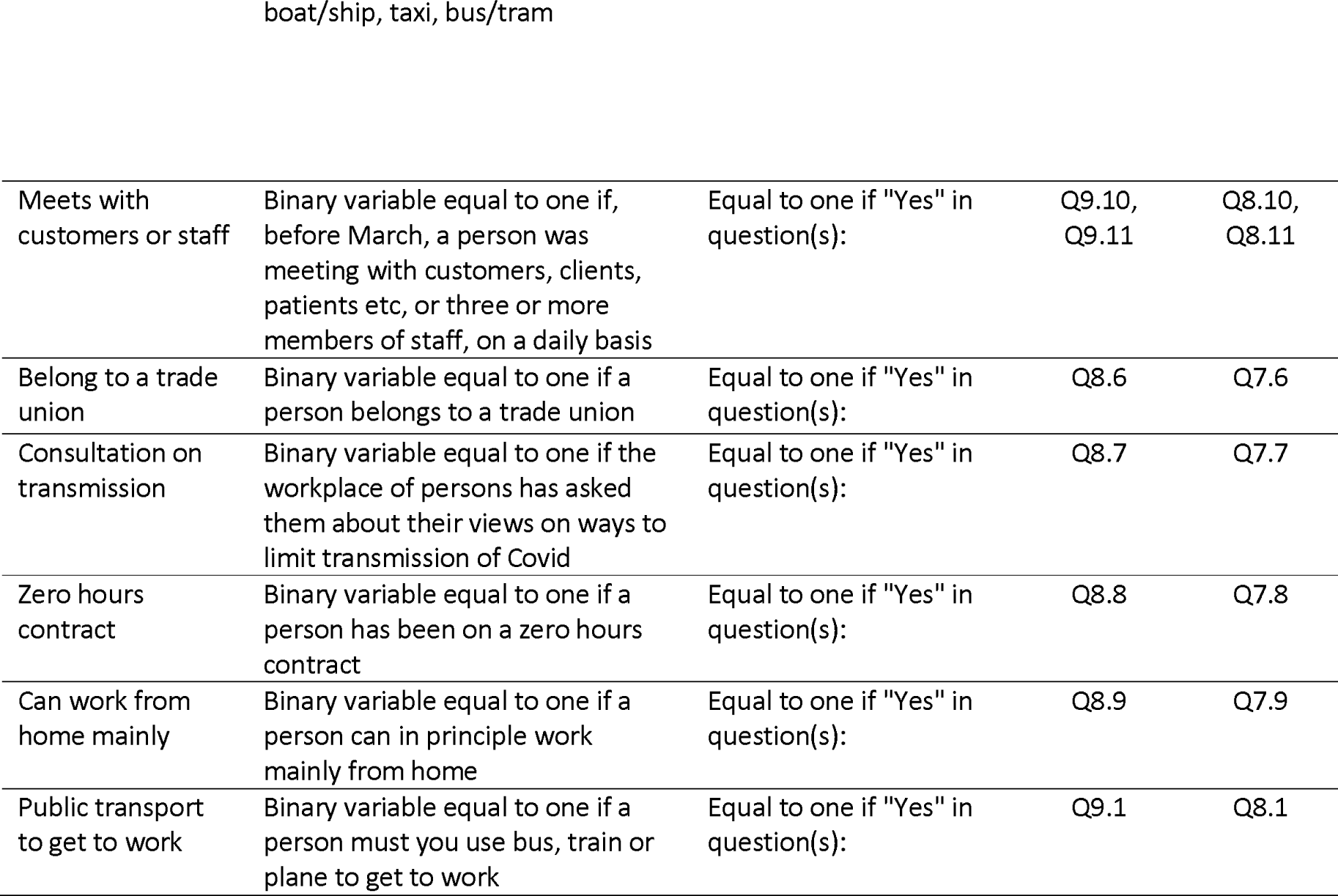

|  | UK |  | US |  | Pool |  |
| --- | --- | --- | --- | --- | --- | --- |
|  | OR | CI | OR | CI | OR | CI |
| **Type of workplace** |  |  |  |  |  |  |
| Airplane | 0.333 | [0.01, 8.18] | 0.167 | [0.01, 5.45] | 0.250 | [0.02, 2.58] |
| Boat/Ship | 3.000 | [0.12, 73.64] | 6.000 | [0.18, 196.28] | 4.000 | [0.39, 41.23] |
| Bus/Tram |  |  | 12.000 | [0.49, 294.57] | 2.000 | [0.22, 17.89] |
| Care-home | 0.207 | [0.02, 2.63] | 0.381 | [0.04, 3.98] | 0.298 | [0.05, 1.64] |
| Factory | 0.462 | [0.04, 5.19] | 0.720 | [0.07, 7.04] | 0.607 | [0.12, 3.15] |
| Food Outlet-Cafe, Takeaway, Restaurant | 0.677 | [0.07, 6.47] | 0.407 | [0.04, 4.25] | 0.545 | [0.11, 2.75] |
| Garden Centre or Farm | 3.000 | [0.2, 45.24] | 1.333 | [0.1, 18.19] | 1.846 | [0.28, 11.98] |
| Hospital | 0.480 | [0.05, 4.67] | 0.943 | [0.1, 8.6] | 0.703 | [0.15, 3.41] |
| Lorry | 2.000 | [0.14, 28.42] | 1.500 | [0.07, 31.57] | 1.800 | [0.25, 12.99] |
| Office | 0.466 | [0.05, 4.03] | 0.232 | [0.03, 2.13] | 0.365 | [0.08, 1.7] |
| Other | 0.144 | [0.01, 1.39] | 0.068 | [0.01, 0.76] | 0.105 | [0.02, 0.54] |
| Prison | 1.500 | [0.07, 31.57] | 1.000 | [0, 0] | 0.600 | [0.05, 7.63] |
| Retail Shop | 0.857 | [0.08, 9.1] | 0.646 | [0.07, 6.17] | 0.710 | [0.14, 3.59] |
| School | 0.462 | [0.05, 4.62] | 0.121 | [0.01, 1.53] | 0.256 | [0.05, 1.37] |
| Taxi | 3.000 | [0.24, 37.67] | 1.333 | [0.1, 18.19] | 2.000 | [0.33, 12.18] |
| Train | 0.462 | [0.02, 8.69] | 1.000 | [0.05, 19.96] | 0.632 | [0.08, 5.1] |
| **Employment status** |  |  |  |  |  |  |
| Working and being paid | 4.179 | [0.97, 17.93] | 4.722 | [0.62, 36.23] | 4.325 | [1.32, 14.13] |
| Working with reduced earnings | 10.339 | [2.43, 44.03] | 15.214 | [2.02, 114.42] | 11.954 | [3.69, 38.71] |
| Redundant or no paid work | 10.977 | [2.46, 48.96] | 15.351 | [2.04, 115.75] | 11.983 | [3.65, 39.39] |
| **Others** |  |  |  |  |  |  |
| Meets with customers or staff | 1.492 | [0.8, 2.78] | 1.234 | [0.68, 2.24] | 1.376 | [0.9, 2.11] |
| Belong to a trade union | 2.102 | [1.25, 3.54] | 7.037 | [4.03, 12.28] | 3.671 | [2.52, 5.35] |
| Consultation about limiting transmission | 2.353 | [1.43, 3.86] | 2.941 | [1.7, 5.07] | 2.602 | [1.8, 3.75] |
| Can work from home mainly | 1.603 | [0.98, 2.63] | 2.380 | [1.38, 4.11] | 1.925 | [1.34, 2.77] |
| Must use public transport to get to work | 1.517 | [0.89, 2.6] | 5.882 | [3.37, 10.26] | 2.843 | [1.94, 4.16] |

| Variable | Definition | Notes on variable construction | US | UK |
| --- | --- | --- | --- | --- |
| Risk preference | Binary variable equal to one if persons self-grade themselves as very prepared to take risks | Equal to one if higher than five in question: | Q14 | Q13 |
| Extraversion | Binary variable equal to one if persons see themselves as extrovert and not reserved | Equal to one if "Yes/Agree" in question(s): | Q8.1 | Q7.1 |
|  |  | and equal to one if "No/disagree" in question(s): | Q8.2 | Q7.2 |
| Frequency of going out | Count variable for visits of more than once a week: a place of work, shops, a place of worship, a cafe, diner, bar, pub or restaurant to eat, drink or take-away food parks, countryside or coast-line, night club or casino, a cinema, museum, concert hall or similar, sports venue, places to entertain children, a family member to provide care or support, social club or bingo | Sum of binary variables equal to "more than once a week" in questions: | Q12.1, Q12.2, Q12.3, Q12.4, Q12.5, Q12.6, Q12.7, Q12.8, Q12.9, Q12.10, Q12.11 | Q11.1, Q11.2, Q11.3, Q11.4, Q11.5, Q11.6, Q11.7, Q11.8, Q11.9, Q11.10, Q11.11 |
| Shared kitchen | Binary variable equal to one if a person shares a kitchen with other households / live in a shared house | Equal to one if "Yes" in question(s): | Q9.6 | Q8.6 |
| University degree | Binary variable equal to one if a person has a university degree in maths, science or economics | Equal to one if "Yes" in question(s): | Q9.8 | Q8.8 |
| Use cash to pay | Binary variable equal to one if a person use cash to pay for things (fares, shopping etc) | Equal to one if "Yes" in question(s): | Q9.7 | Q8.7 |
| Own a car | Binary variable equal to one if a household owns a car | Equal to one if "Yes" in question(s): | Q9.2 | Q8.2 |
| Not able to keep social distance | Variable equal to zero if persons are able to keep social distance at their local neighborhood outside their house, or when shopping; variable to -1 in other cases. | Equal to -1 when the answers are "Not applicable to me" or "Sometimes / never" in questions: | Q5.3, Q5.4 | Q4.3, Q4.4 |
| Live with others | Binary variable equal to one if a person lives with their parents, children or parents | Equal to one if "Yes" in question(s): | Q9.3, Q9.4, Q9.5 | Q8.3, Q8.4, Q8.5 |
| Spend time at home with others | Binary variable equal to one if a person spends time with others when at home | Equal to one if "Yes" in question(s): | Q8.5 | Q7.5 |
| Residential Area | Binary variable equal to one if a person lives in a city | Equal to one if a person lives in a high rise apartment/flat or other in a city in question: | Q11 | Q10 |
| Type of workplace | Binary variable equal to one if a person works in an airplane, care-home, factory, food, hospital, office, retail shop or school (protective workplace). Zero in transport-related workplaces: | Classifications based on question "Which of these best describes your main current workplace?" | Q10 | Q9 |

|  |  |  |  |  |
| --- | --- | --- | --- | --- |
| Meets with customers or staff | Binary variable equal to one if, before March, a person was meeting with customers, clients, patients etc, or three or more members of staff, on a daily basis | Equal to one if "Yes" in question(s): | Q9.10, Q9.11 | Q8.10, Q8.11 |
| Belong to a trade union | Binary variable equal to one if a person belongs to a trade union | Equal to one if "Yes" in question(s): | Q8.6 | Q7.6 |
| Consultation on transmission | Binary variable equal to one if the workplace of persons has asked them about their views on ways to limit transmission of Covid | Equal to one if "Yes" in question(s): | Q8.7 | Q7.7 |
| Zero hours contract | Binary variable equal to one if a person has been on a zero hours contract | Equal to one if "Yes" in question(s): | Q8.8 | Q7.8 |
| Can work from home mainly | Binary variable equal to one if a person can in principle work mainly from home | Equal to one if "Yes" in question(s): | Q8.9 | Q7.9 |
| Public transport to get to work | Binary variable equal to one if a person must use bus, train or plane to get to work | Equal to one if "Yes" in question(s): | Q9.1 | Q8.1 |

## Footnotes

1 Ethics approval was granted by the ethics review board under HREC/3590.

